# ChatGPT 4 Versus ChatGPT 3.5 on The Final FRCR Part A Sample Questions. Assessing Performance and Accuracy of Explanations

**DOI:** 10.1101/2023.09.06.23295144

**Authors:** Youssef Ghosn, Omar El Sardouk, Yara Jabbour, Manal Jrad, Mohammed Hussein Kamareddine, Nada Abbas, Charbel Saade, Alain Abi Ghanem

## Abstract

**Objective:** To evaluate the performance of two versions of ChatGPT, GPT4 and GPT3.5, on the Final FRCR (Part A) also referred to as FRCR Part 2A radiology exam. The primary objective is to assess whether these large language models (LLMs) can effectively answer radiology test questions while providing accurate explanations for the answers.

**Methods:** The evaluation involves a total of 281 multiple choice questions, combining the 41 FRCR sample questions found on The Royal Collage of Radiologists website and 240 questions from a supplementary test bank. Both GPT4 and GPT3.5 were given the 281 questions with the answer choices, and their responses were assessed for correctness and accuracy of the explanations provided. The 41 FRCR sample questions difficulty was ranked into “low order” and “high order” questions. A significance level of p<0.05 was used.

**Results:** GPT4 demonstrated significant improvement over GPT3.5 in answering the 281 questions, achieving 76.5% correct answers compared to 52.7%, respectively (p<0.001). GPT4 demonstrated significant improvement over GPT3.5 in providing accurate explanations for the 41 FRCR sample questions, with an accuracy of 65.9% and 31.7% respectively (p=0.002). The difficulty of the question did not significantly affect the models’ performances.

**Conclusion:** The findings of this study demonstrate a significant improvement in the performance of GPT4 compared to GPT3.5 on FRCR style examination. However, the accuracy of the provided explanations might limit the models’ reliability as learning tools.

**Advances in Knowledge:** The study indirectly explores the potential of LLMs to contribute to the diagnostic accuracy and efficiency of medical imaging while raising questions about the current LLMs limitations in providing reliable explanations for radiology related questions hindering its uses for learning and in clinical practice.

**Highlights:**

- ChatGPT4 passed an FRCR part 2A style exam while ChatGPT3.5 did not.
- ChatGPT4 showed significantly higher correctness of answers and accuracy of explanations.
- No significant difference in performance was observed between “high order” and “lower order” questions.
- Explanation accuracy was lower than correct answers rate limiting the Models’ reliability as
- learning tools.

## Introduction

Throughout the 21st century, artificial intelligence (AI) has seen exponential growth in its capabilities and potential, leading to increased interest in what it can provide to each respective field. ChatGPT, short for Chat Generative Pre-trained Transformer and referred to simply as GPT in this study, is an advanced language model developed by OpenAI. GPT 4 and its predecessor GPT3.5 are large language models (LLM) capable of processing image and text inputs as well as producing text outputs (1). GPT 4 has been evaluated using several standardized exams in comparison to GPT 3.5, such as the Uniform Bar Exam, Law School Admission Test (LSAT), SAT math, and AP psychology to name a few. It passed all these exams with generally high scores and had considerably improved compared to its predecessor GPT 3.5 (1). A study was done by Kung, T.H. et al to evaluate the performance of GPT 3.5 (GPT 4 was not developed at the time of the study) on the USMLE step 1, step 2CK, and step 3 exams. These exams generally require extensive medical knowledge along with correlating findings in the given vignette to find the correct answer or exclude incorrect options. The study concluded that GPT 3.5 yields moderate accuracy approaching passing performance on the USMLE exams (2). A previous study done by Bhayana, R. et al in early 2023 evaluated GPT 3.5 on the Canadian Royal College and American Board of Radiology examinations with 150 multiple-choice questions similar in their style, content, and difficulty (3). The results revealed that GPT 3.5 answered 69% (104 out of 150) of the questions correctly, nearly passing the radiology board-style examination [9]. The study managed to highlight the potential of ChatGPT as a useful tool for radiologists. However, as GPT 4 had not been developed at the time of the study, it would be interesting to see how far AI has come in realizing its potential in radiology.

A study evaluating the performance of GPT 4/3.5 on the final Fellowship of the Royal College of Radiologists (FRCR) Part 2A, or even GPT 4 on any radiological examination, has yet to be done. The FRCR Part 2A is a standardized examination that assesses the knowledge of radiology trainees regarding pathology, imaging techniques, congenital abnormalities, and radiological findings that underpin the practice of radiology. It is the final written examination in general clinical radiology and is expected to be completed during core clinical training. It is made of two exam papers covering all the radiological subspecialties, with each paper consisting of 120 questions with a single best answer. Examinees are given 3 hours to finish each paper. The prospect of an AI model passing the FRCR Part 2A would suggest that AI can play a part in the evolution of the profession of radiology and further raise interest in further radiology-related AI research (4,5). Moreover, if the explanations given by GPT are accurate, GPT might be used as an adjunct reliable learning tool. In this paper we assess whether GPT3.5 and GPT4 can correctly answer the FRCR Part 2A test style questions of various difficulty while providing accurate explanations.

## Methods

To evaluate the performance of GPT 4/3.5 on the FRCR part 2A, 41 questions from the FRCR sample provided online on the Royal Collage of Radiologist (RCR) official website were used (4,6) along with 240 questions covering all modules from a test bank to supplement them (7), for a total of 281 questions. It is important to mention that the questions do not include any images, and therefore, will not assess GPT 4/3.5’s ability to read radiological imaging or hinder its ability to answer questions that do include them. The revised Bloom Taxonomy for learning and assessment in radiology was used to classify the questions into either “low-order” (knowledge, comprehension, and application” or “high-order” (analysis, synthesis, and evaluation) thinking questions (8).

The questions with the answer choices were plugged in GPT 4/3.5 and require of it to choose the correct answer with justification. No prompts were provided. Using the provided answer key as well as the acquired knowledge and experience, one PGY5 (final year) and one PGY 4 radiologists in training at the American University of Beirut determined the accuracy of GPT 4/3.5’s responses. Accuracy was defined as both choosing the correct answer and the correct justification/explanation. Incorrect answers and correct answers with false justification/explanation were considered inaccurate. The two radiologists analyzed independently, then shared their findings to discuss them and reach a consensus.

### Statistical analysis

The study compared performance and accuracy by question types (low order vs. high order), by model (GPT4 vs GPT3), and by exam (FRCR sample vs test bank) using the Chi-square test and Fisher exact test. A significance level of p<0.05 was used to determine significant differences. All statistical analyzes were performed using SPSS.

## Results

GPT 3.5 managed to have 52.7% of questions correctly (N= 148) of the 281 questions, which include both the test bank and the FRCR sample. Meanwhile, GPT 4 had 76.5% of the questions answered correctly (N=215), which was significantly better than its predecessor GPT 3.5 (p< 0.001) (Table 1, figure 1).

**Table 1:** Performance and accuracy of Chat-GPT3.5 vs Chat-GPT 4 on FRCR sample and test bank sample.

|  | Chat-GPT-3.5 |  | Chat-GPT-4 |  |  |
| --- | --- | --- | --- | --- | --- |
|  | n | % | n | % | Significance,<br>Chi-square<br>test |
| **Answer (Test bank + FRCR sample)** |  |  |  |  | **<0.001\*** |
| correct | 148 | 52.7% | 215 | 76.5% |  |
| Incorrect | 133 | 47.3% | 66 | 23.5% |  |
| Total | 281 | 100% | 281 | 100% |  |
| **Answer (FRCR sample)** |  |  |  |  | **0.0051\*** |
| Correct | 20 | 48.8% | 33 | 80.5% |  |
| Incorrect | 21 | 51.2% | 8 | 19.5% |  |
| Total | 41 | 100% | 41 | 100% |  |
| **Accuracy (FRCR sample)** |  |  |  |  | **0.002\*** |
| Accurate | 13 | 31.7% | 27 | 65.9% |  |
| Inaccurate | 28 | 68.3% | 14 | 34.1% |  |
| Total | 41 | 100% | 41 | 100% |  |

\*Indicates statistically significant finding.

**Figure 1:**
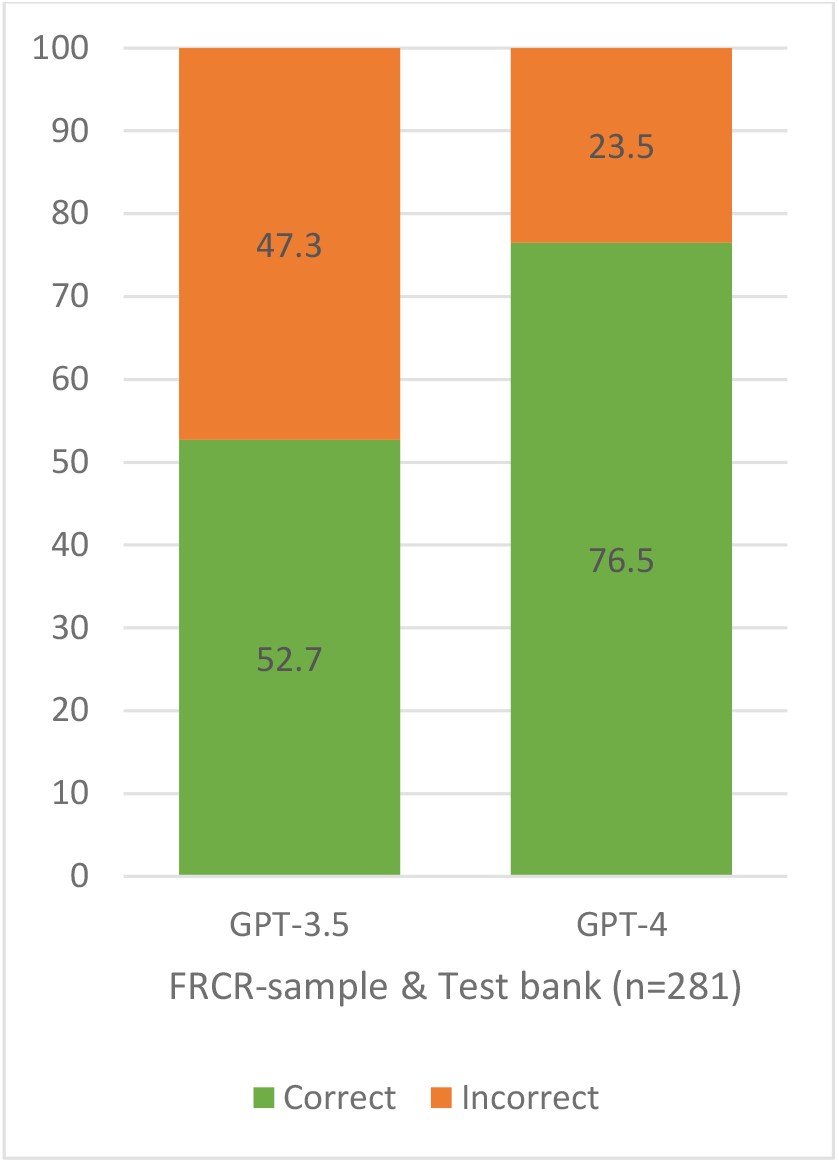
Percentage of correct and incorrect answers. GPT-4 versus GPT 3.5

When analyzing the accuracy of explanations for both LLMs (Table 1), only the FRCR sample (N=41) was used. It was found that GPT 3.5 and 4 had a score of 20 (48.8%) and 33 (80.5%) correct answers, respectively (p=0.0051). Moreover, of the 20 correct answers, 13 answers (65%) were found to be accurate in GPT 3.5; whereas, of the 33 correct answers, 27 answers (81.8%) were found to be accurate in GPT 4. The total accuracy of GPT 4 was 65.9% which was significantly greater than GPT 3.5 being 31.7% (p=0.002).

When observing whether there was a difference in performance between the test bank and the FRCR sample with both LLMs (table 2, figure 1), there was no noticeable difference to be found; GPT 3.5 had 53.3% and 48.8% correct, respectively (p=0.589); meanwhile, GPT 4 had 75.8% and 80.5%, respectively (p=0.516).

**Table 2:** Performance of ChatGPT on FRCR sample and test bank sample.

|  | FRCR-samples |  | Test bank<br>Sample |  |  |
| --- | --- | --- | --- | --- | --- |
|  | n | % | n | % | Significance,<br>Chi-square<br>test |
| **ChatGPT-3.5** |  |  |  |  | 0.589 |
| Correct | 20 | 48.8% | 128 | 53.3% |  |
| Incorrect | 21 | 51.2% | 112 | 46.7% |  |
| Total | 41 | 100% | 240 | 100% |  |
| **ChatGPT-4** |  |  |  |  | 0.516 |
| Correct | 33 | 80.5% | 182 | 75.8% |  |
| Incorrect | 8 | 19.5% | 58 | 24.2% |  |
| Total | 41 | 100% | 240 | 100% |  |

Potential differences in performance on “low-order” and “high-order” FRCR part 2A questions were analyzed using only the online FRCR sample (table 3, figure 2). Nineteen questions were classified as “high-order”, while 22 questions were classified as “low-order”. GPT 4 managed to answer 15 (78.9%) “high-order” and 18 (81.1%) “low-order” questions correctly; whereas, GPT 3.5 had 8 (42.1%) “high-order” and 12 (42.5%) “low-order” questions correct. When observing whether the performance of GPT 4 and 3.5 differed between “high-order” and “low-order” questions, no significant changes could be found with both LLMs (p=0.817 and p=0.427, respectively).

**Table 3:**
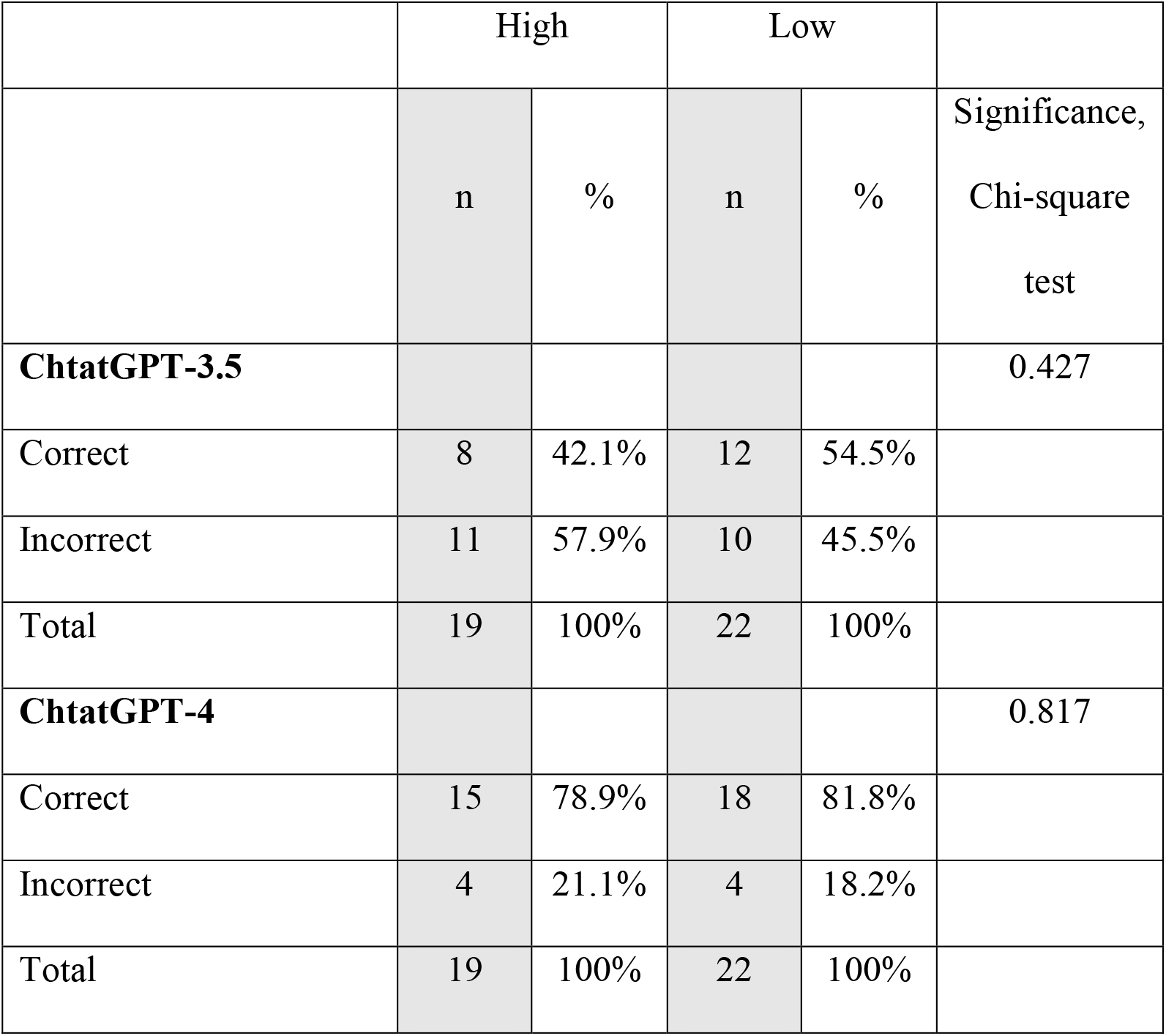
ChatGPT-3.5 and ChatGPT-4 performance depending on the difficulty of the question on the FRCR sample.

**Figure 2:**
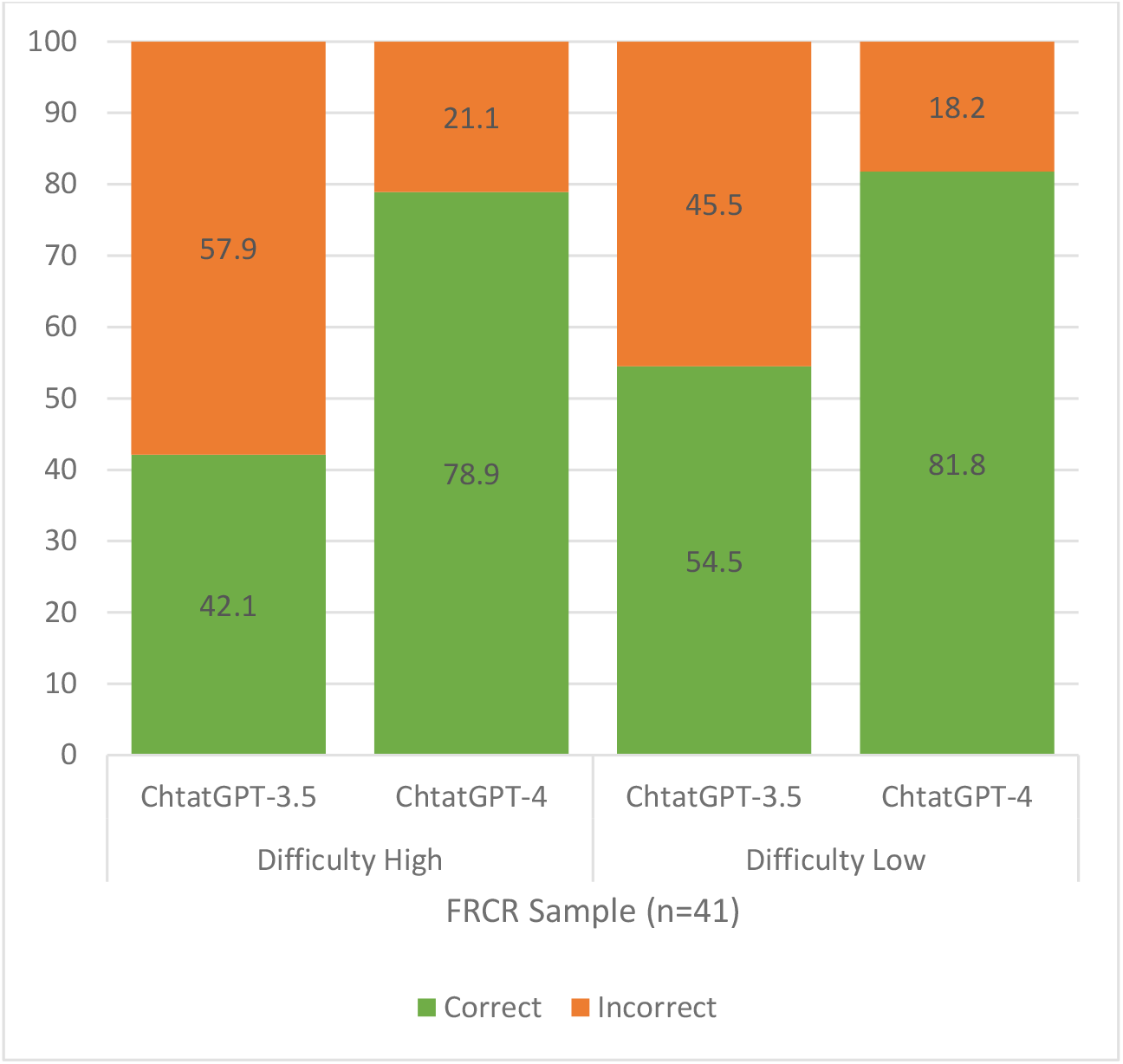
Percentage of correct and incorrect Chat-GPT answers for low- and high-order questions.

Furthermore, GPT 4 had 11 (73.3%) “high-order” and 16 (88.8%) “low-order” correct questions that were found to be accurate, while, GPT 3.5 had 5 (62.5%) “high-order” and 8 (66.6%) “low-order” correct questions that were found to be accurate (tables 4 and 5). Both models had more accurate explanations for “low-order” questions compared to “high-order” questions. However, accuracy was not found to be statistically different when comparing high- and lower-order questions (p=0.375 for GPT4 and p=0.999 for GPT3.5).

**Table 4:**
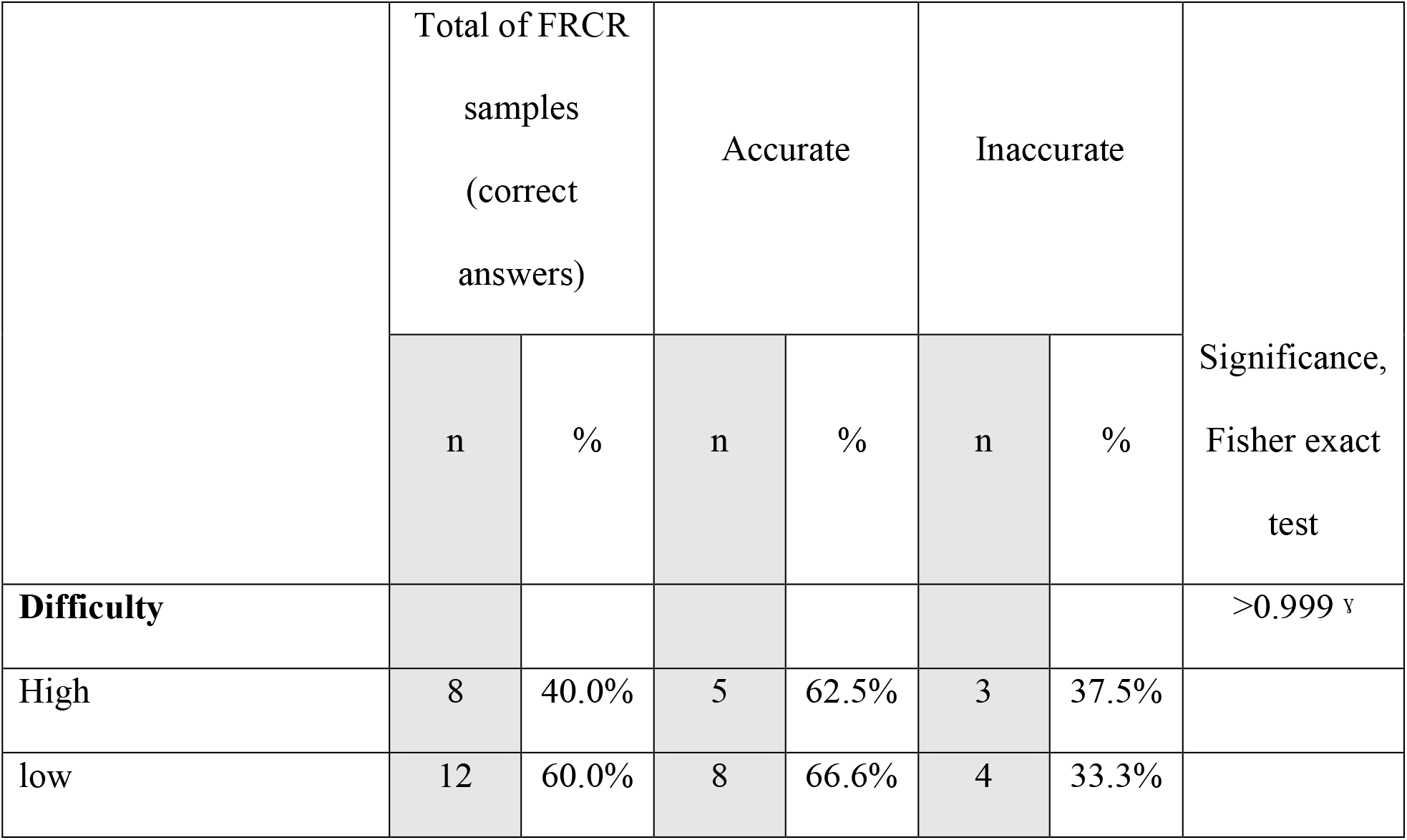
for Chat-GPT-3.5, comparing difficulty and accuracy on correct answers of FCRC answers.

|  | Total of FRCR<br>samples<br>(correct<br>answers) |  | Accurate |  | Inaccurate |  | Significance,<br>Fisher exact<br>test |
| --- | --- | --- | --- | --- | --- | --- | --- |
|  | n | % | n | % | n | % |  |
| **Difficulty** |  |  |  |  |  |  | >0.999 <sup>y</sup> |
| High | 8 | 40.0% | 5 | 62.5% | 3 | 37.5% |  |
| low | 12 | 60.0% | 8 | 66.6% | 4 | 33.3% |  |

**Table 5:** for Chat-GPT-4, comparing difficulty and accuracy on correct answers of FCRC answers.

|  | Total of FRCR<br>samples<br>(correct<br>answers) |  | Accurate |  | Inaccurate |  | Significance,<br>Fisher exact<br>test |
| --- | --- | --- | --- | --- | --- | --- | --- |
|  | n | % | n | % | n | % |  |
| **Difficulty** |  |  |  |  |  |  | 0.375 <sup>y</sup> |
| High | 15 | 45.5% | 11 | 73.3% | 4 | 26.6% |  |
| low | 18 | 54.5% | 16 | 88.8% | 2 | 11.1% |  |

Finally, the total accuracy of GPT4 and GPT3.5 for the 41-question sample was 65.9% and 31.7%, respectively (Figure 3).

**Figure 3:**
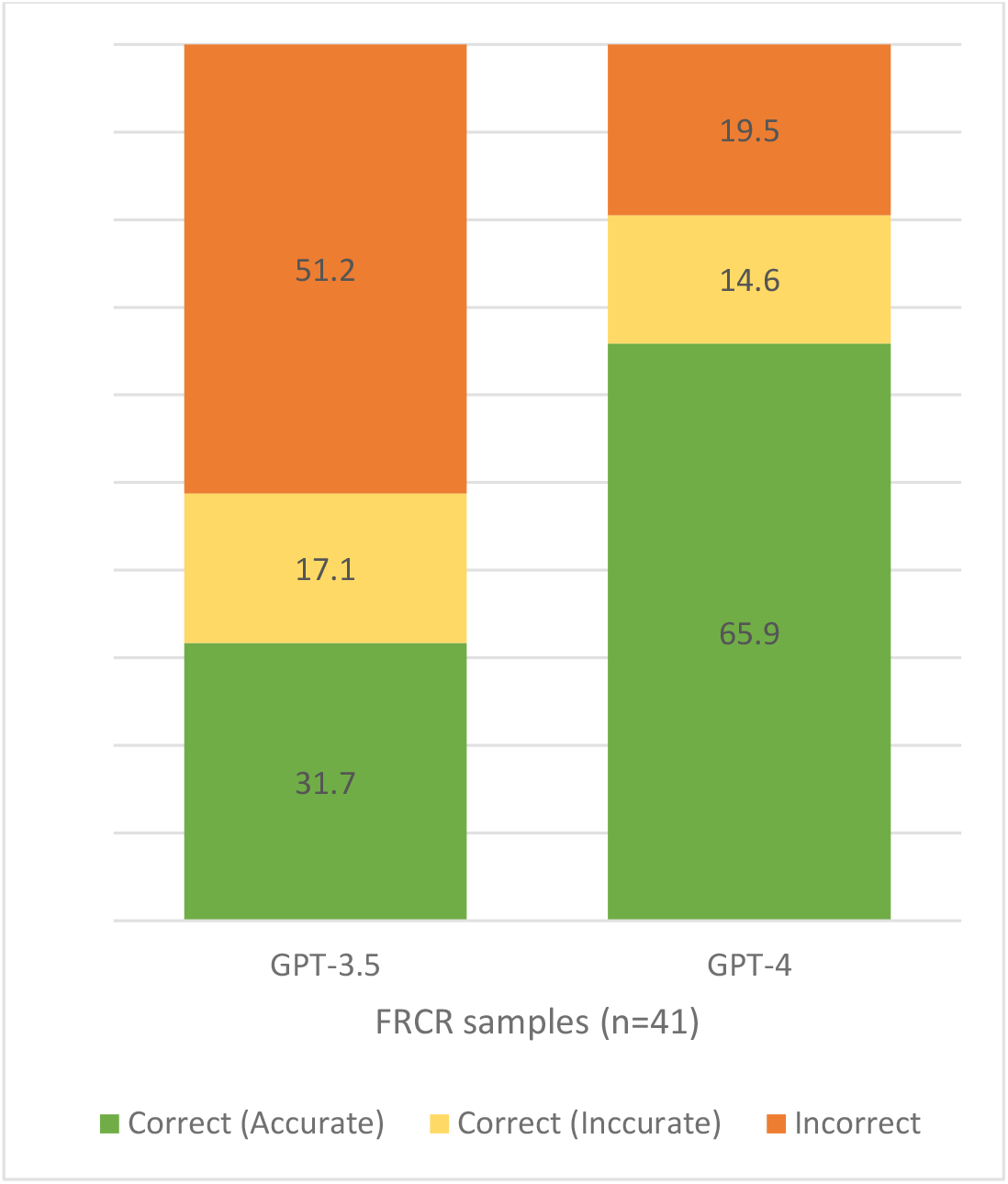
total accuracy of explanations of GPT-3.5 and GPT-4.

## Discussion

The analysis of GPT 3.5 and GPT 4 in the FRCR 2A radiology exam provides valuable insight into how these language models could function in a clinical radiology setting. The results demonstrate the potential of AI models to aid in decision-making and increase interest in further research into the role of AI in radiology.

Compared to GPT 3.5, GPT 4 showed a significant improvement in performance on the FRCR 2A test. GPT 4 answered 76.5% of both test banks’ questions correctly, while GPT 3.5 had 52.7% of its answers correct. All the while, GPT 4 had an accuracy of 65.9%, significantly exceeding GPT 3.5’s 31.7% accuracy rate. This finding indicates that improvements in the multipotent GPT 4 have contributed to its performance enhancement. These results are consistent with previous studies that have shown better performance of GPT 4 compared to its predecessors in controlled trials (1). Assuming a 60% passing rate cutoff for the FRCR Part 2A exam, GPT 4 managed to pass the exam with a rate of 76.5%; meanwhile, GPT 3.5 did not.

In addition, the study examined performance on GPT 3.5 and GPT 4 on “higher order” and “lower order” questions. Surprisingly, neither model showed a significant difference in performance between these questions. Both models exhibited similar levels of accuracy for the two groups, indicating that the level of difficulty did not affect the model’s ability to answer questions. However, GPT-4 consistently outperformed GPT-3.5 in terms of overall correctness and accuracy, demonstrating improved performance on both “high” and “low” order-rated questions.

It is important to note that GPT 4 demonstrated an explanation accuracy rate of 65.9%, dropping from the correct rate of 80.5%. This highlights ChatGPT’s shortcomings in providing meaningful explanations for the answers, ultimately limiting its reliability as a learning tool.

Regardless, this is a substantial improvement. The notion that an AI algorithm with no radiology training can pass a standardized examination in radiology, especially when its evolutionary predecessor could not, suggests that we will soon enough reach an era where AI, possibly the GPT lineage, will reliably serve as a tool for radiologists to benefit from as mentioned before. With the incorporation of imaging processing systems along with continued improvement in language processing, specifically the development of generalist foundation models for radiology and medical AI, there remains a vast amount of untapped potential (9,10).

Figure 4 and Figure 5 present two questions from the FRCR online sample used. The first question was answered correctly by GPT3.5, however, the provided explanation was inaccurate. The second case tests radiologists’ ability to suspect potential child abuse. GPT3.5 and GPT4 failed to answer the question correctly, highlighting GPT’s potential weakness in processing high order questions with ethical/societal concerns at hand.

**Figure 4.**
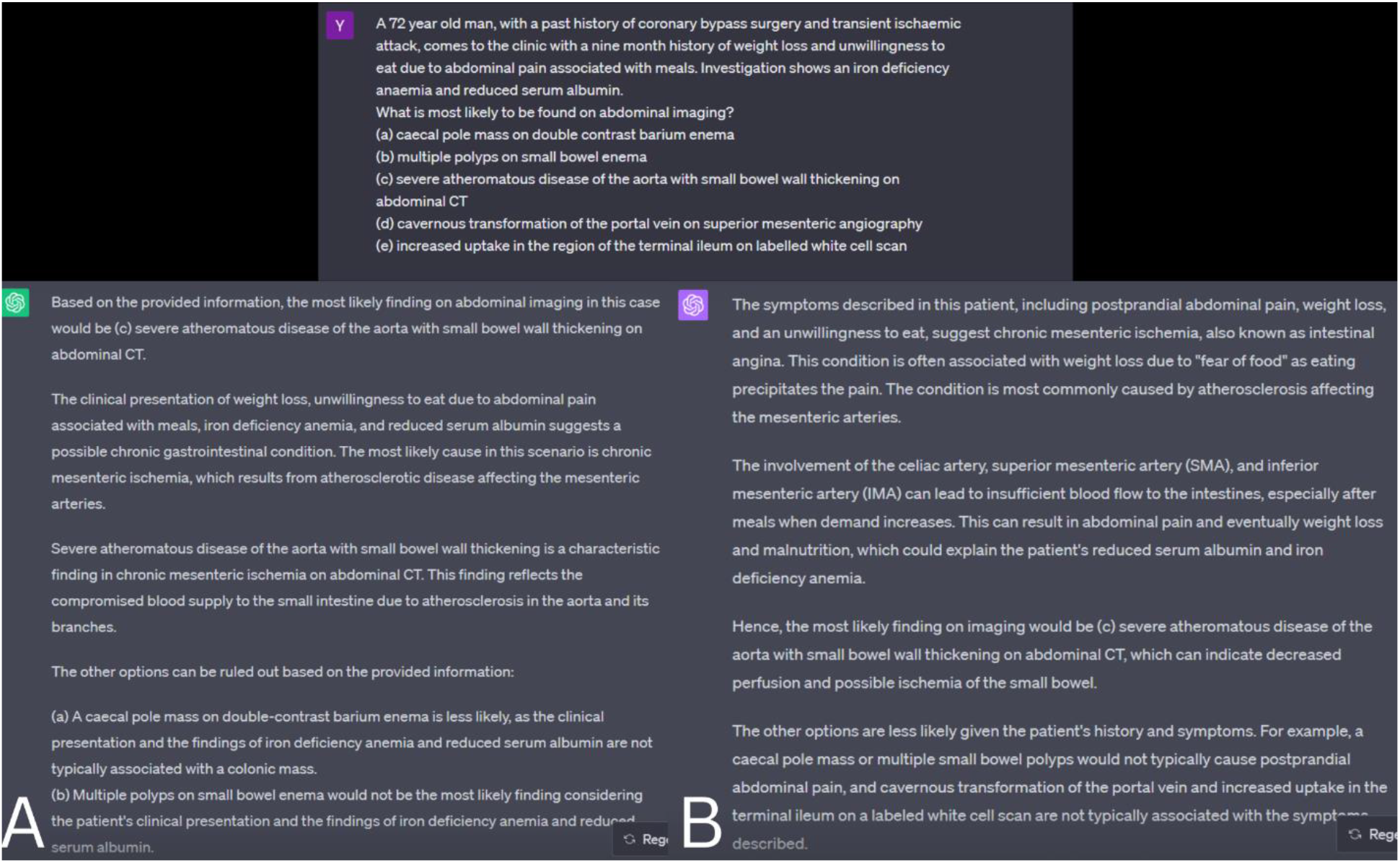
GPT3.5 (A) correctly answers the question identifying the diagnosis of chronic mesenteric ischemia and inferring the suspected image finding as “severe atheromatous disease of the aorta with small bowel wall thickening on abdominal CT”. However, in the explanation the model ruled out a cecal pole mass explaining that “the findings of iron deficiency anemia and reduced serum albumin not typically associated with a colonic mass” which is an inaccurate explanation as iron deficiency anemia is a common symptom in patients with colorectal cancer. GPT4 (B) did not proved factually incorrect statements in the explanation.

**Figure 5.**
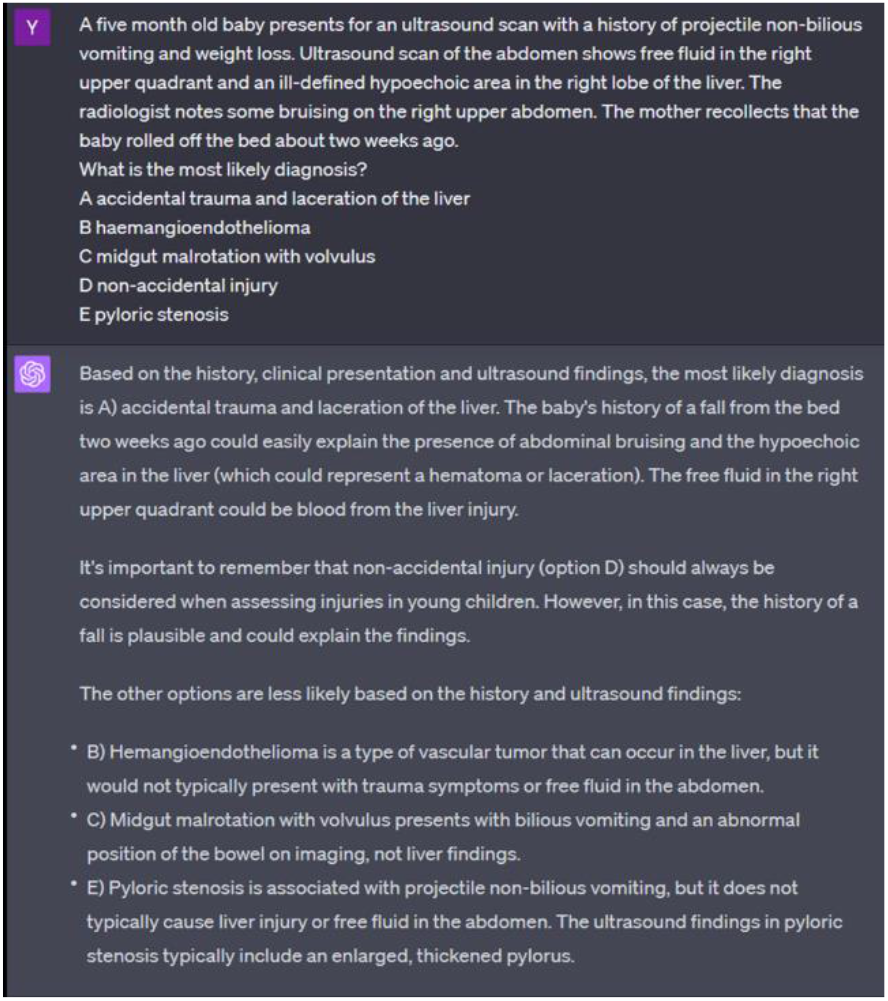
GPT4 failed to correctly answer a diagnosis of non-accidental injury. The model here points out that non-accidental injury should always be considered when evaluating trauma in young children, however, it considered the history of a fall as a plausible explanation for accidental trauma. The model failed to detect the mismatch between the clinical findings and the inconsistent history provided by the mother. An accidental fall over a short distance (from the bed), not prompting the mother to seek medical attention for 2 weeks, is unlikely to account for the liver laceration, free fluid, and abdominal bruising. This raises the stakes for a non-accidental injury which should be considered first (4).

## Limitations

The study was based solely on text-based questionnaires and did not consider the interpretability of images. The inclusion of image-based questionnaires may lead to a moving analysis beyond simple performance in radiological observations. Another limitation is the small sample size of the FRCR Part 2A questions available, as the questions are confidential and out of reach. This, coupled with the fact that the questions supplemented from the test bank have not been analyzed for accuracy or difficulty, may limit the generalizability of the results due to the small sample size. Finally the absence of prompts might have affected the models answers.

## Conclusion

The findings of this study demonstrate a significant improvement in the performance of ChatGPT 4 compared to ChatGPT 3.5 on radiology FRCR part 2A style examination. Although GPT 4 showed promising results in the FRCR trial, the accuracy of the provided explanations limits its reliability as a learning tool. Finally, further research is needed to evaluate its performance on image-based questions and explore its potential in real-world clinical settings, acting as a complementary tool for human knowledge.

## Data Availability

All data produced in the present study are available upon reasonable request to the authors

## Abbreviations

FRCR: Fellowship of Royal College of Radiologists
RCR: Royal College of Radiologists
LMLs: Large Language Models
ChatGPT: Chat Generative Pre-trained Transformer
AI: Artificial Intelligence
LSAT: Law School Admission Test
USMLE: United States Medical License Examination
PGY: Post Graduate Year

## References

1. OpenAI. GPT-4 Technical Report. 2023;4:1–100. Available from: http://arxiv.org/abs/2303.08774

2. Kung TH, Cheatham M, Medenilla A, Sillos C, De Leon L, Elepaño C, et al. Performance of ChatGPT on USMLE: Potential for AI-assisted medical education using large language models. PLOS Digit Heal. 2023;2(2):e0000198.

3. Bhayana R, Krishna S, Bleakney RR. Performance of ChatGPT on a Radiology Board-style Examination: Insights into Current Strengths and Limitations. Radiology. 2023;307(5).

4. The Royal Collage of Radiologists. Final FRCR Part A – Guidance for Candidates. :1–32. Available from: https://www.rcr.ac.uk/sites/default/files/how_to_approach_cr2a_candidate_guidance.pdf

5. The Royal Collage of Radiologists. Final FRCR Part B Examination - Purpose of Assessment Statement. Available from: https://www.rcr.ac.uk/sites/default/files/cr2b_purpose_of_assessment_statement.pdf

6. The Royal Collage of Radiologists. Final Examination for the Fellowship in Clinical Radiology (Part B) Guidance Notes for Candidates. 2015;(c). Available from: https://www.rcr.ac.uk/sites/default/files/cr2a_guidance_notes._jan_22.pdf

7. Teck Yew Chin, Akash Ganguly CA. Get Through Final FRCR 2A [Internet]. 1st ed. Get Through Final FRCR 2A. Taylor & Francis; 2017. Available from: https://www.amazon.com/Get-Through-Final-FRCR-2A/dp/1138743992

8. Smith EB, Gellatly M, Schwartz CJ, Jordan S. Training Radiology Residents, Bloom Style. Acad Radiol. 2021;28(11):1626–30.

9. Wu C, Zhang X, Zhang Y, Wang Y, Xie W. Towards Generalist Foundation Model for Radiology. 2023;1–24. Available from: http://arxiv.org/abs/2308.02463

10. Moor M, Banerjee O, Abad ZSH, Krumholz HM, Leskovec J, Topol EJ, et al. Foundation models for generalist medical artificial intelligence. Nature. 2023;616(7956):259–65.

